# Assessment of Working Environment and Satisfaction Among Health Care Workers In Sengerema District Mwanza, Tanzania

**DOI:** 10.1101/2023.05.15.23290008

**Authors:** Peter Chilipweli, Namanya Basinda, Evans V Msilaga, Tusherahma Tuli, Frank setebe, Emanuel Elias, Eveline Konje, Anthony Kapesa, Elias Nyanza, Dominica Morona

**Affiliations:** Department of Community Medicine, School of Public Health, Catholic University of Health and Allied Sciences (CUHAS), Box 1464, Mwanza; Department of Biostatistics and Epidemiology, School of Public Health, Catholic University of Health and Allied Sciences (CUHAS), Box 1464, Mwanza; Department of Medical Parasitology and Entomology, School of Public Health, Catholic University of Health and Allied Sciences (CUHAS), Box 1464, Mwanza; Department of environmental occupational health and GIS, School of Public Health, Catholic University of Health and Allied Sciences (CUHAS), Box 1464, Mwanza; Weill bugando school of medicine, Catholic University of Health and Allied Sciences (CUHAS), Box 1464, Mwanza

**Keywords:** Job Sastification, health care workers, dispensary, Health center, Job turnover, quality improvement, Sengerema

## Abstract

**Background:** Job satisfaction is currently considered to be a measure that should be included in quality improvement programs thus it is very important to assess it among healthcare workers who are mostly exposed to work stress thus this research is going to have different consideration on health care workers working on hard-to-reach areas and low-level facilities since there is lack of enough evidence and limited studies done under this area.

**Methods:** This was a cross section study conducted among healthcare workers found in sengerema and it was conducted from 27/02/2023 to 10/03/2023 whereby it include 356 health care workers employed and working at sengerema district as participants found in the health facilities Statistical method used was descriptive analysis by assessing frequency of variables in terms percentages. The results were presented in form of tables and bar graphs.

**Results:** A total of 356 health care professionals were included in the study, female participants were more occupying 55.6% while the male were 44.4%. In the study more of the participants were married than those single with percentages of 68.5% and 31.5% respectively. Most of the health care workers were at the dispensary level were satisfied at their job (55%0 but among the health care workers working at the health center most of them were not satisfied (52%). But among married workers most of them were not satisfied with their job (50.5%) as compared to single workers who were most satisfied (56%).

**Conclusion:** Based on our research, the prevalence of job satisfaction among health care workers in Sengerema district is generally high by 53.9% which shows that most workers are satisfied with their jobs. Most health care workers have good relationship with each other and can depend on each other and they get their benefit as health care workers.

## Introduction

Job satisfaction is currently considered to be a measure that should be included in quality improvement programs. In health care organizations, it is very essential to determine factors associated with job satisfaction since this will ensure the provision of quality of care, as well as organizational efficiency, and effectiveness. Work place environment impact the morale, productivity engagement and job satisfaction both positively and negatively.

Montaged and colleagues in their study showed that the dissatisfaction of the health care workers can be due to long working hours, working environment conditions, and weakness in the way of reward and punishment and evaluation method (8). Adams and Bond in their study concluded that organizational factors are more important than individual factors in the prediction of the health care workers job satisfaction (9). Lu Research is also suggesting that factors such as education level, shifts and tasks can be effective on job satisfaction (10). Stress among health care workers is considered as a major problem worldwide.

A study on work satisfaction of professional HCW in South Africa by Pillay 2008 indicated overall dissatisfaction among South African HCW and highlighted the disparity between levels of job satisfaction in the public and private sectors. In Tanzania, studies provide strong evidence for the clinical efficacy and economic value of these cadres, particularly in the provision of emergency obstetric care. Given these indicators, it is important to recruit, retain and support these cadres to build the capacity of health systems in low-income countries.

Research done in Dar es salam by Naburi et al. on job satisfaction and turnover intention among health care stuffs providing services for prevention of mother to child transmission of HIV shows that 54% of the providers were dissatisfied with their current job due to low salaries and high work load, but satisfied with work place harmony and being able to follow their moral values (14) Also, Research done by Maryam Ukwaju (SAUT, 2012) on the effects of employee’s job satisfaction on work performance at Mwanza city council shows high percent of employee are not satisfied with pay and opportunities for promotion to a large extent are not motivated by them too. Factors like working condition, job security, supervision and leadership, organization policy technology change and organization culture affects employee’s performance. (15) While the study done by Leshabari (2008) focused on motivation and factors associated with low motivation, this study addressed the issue of level of job satisfaction among HCW, as well as the determinants of the different degrees of job satisfaction among health care workers (16). A research done in Dar es Salaam by Naburi et al. On job satisfaction and turnover intention among health care stuffs providing health services for prevention of mother to child transmission of HIV. Therefore, this research is going to have different consideration on health care workers working on hard-to-reach areas and low-level facilities since there is lack of enough evidence and limited studies done under this area. Therefore, it has assessed job satisfaction among H.C.W in Sengerema district and determining job relation and benefits among H.C.W in Sengerema district. Whereby findings from this study provide understandings on working environment and level of satisfaction among health care worker in public health facilities at Sengerema district in Mwanza Tanzania. Healthcare stakeholders are expected to benefit from the findings by understanding the impact of working environment and job satisfaction among health care workers in Tanzania health facilities.

## METHOD

### Study area

This study was conducted at Sengerema district in Mwanza region. It is bordered to the north and east by Lake Victoria, to the south by Geita Region and to the southeast by the Misungwi District. There are 48 health facilities in Sengerema district.

**Figure 1:**
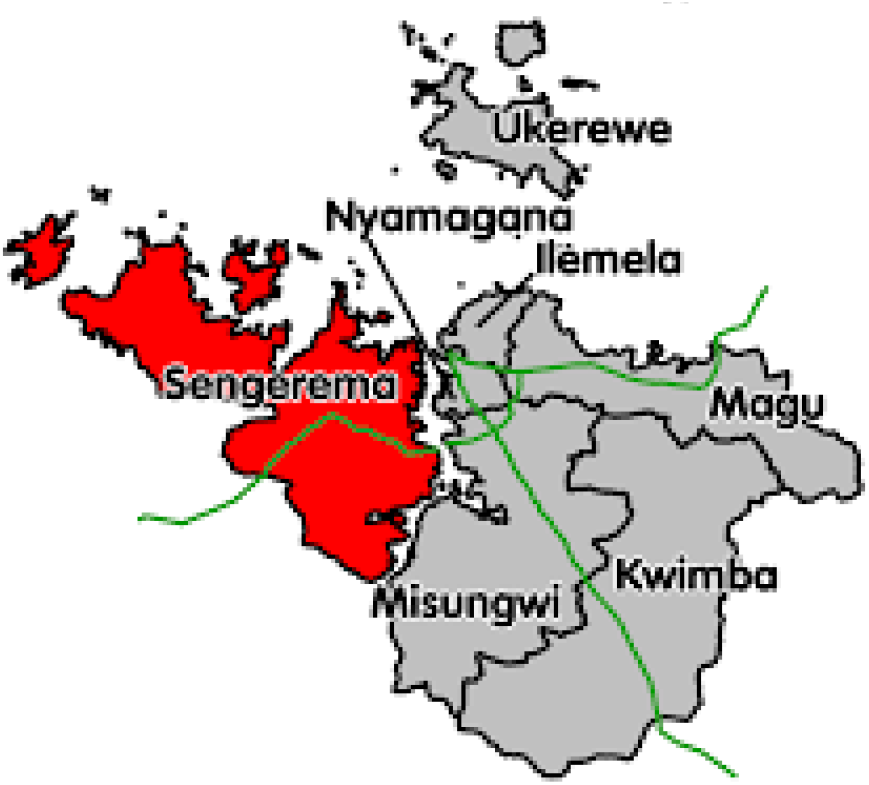
Map of sengerema district

#### Study design

This was a cross section study conducted among medical practitioners found in sengerema and it was conducted from 27/02/2023 to 10/03/2023, because of short duration for data collection hence it was seen to suite the purpose, since it is Easy and quick, less costly and allows to compare many different variables at the same time.

### Study population

This study included health care workers employed 366 participants found in the health facilities of Sengerema district.

## SELECTION CRITERIA

### Inclusion criteria

HCWs who were present and working in the health facility

### Exclusion criteria

HCWs who have less than 3 months at work and those who were on special program at the health facility by the time of data collection.

### Sample size and sampling procedure

The study involved 356 Health care workers which is the coverage of the study population in Sengerema district. Therefore, sample size estimation was not performed and no sampling procedure was employed since all health care workers were involved

### Data collection

The data collection was done in form of questionnaire which was semi structured and close ended. The questionnaire was adopted from CDC, then translated to Swahili language form because it is the national language of the country.

### Data analysis

After data collection, data was entered in the Microsoft excel and analyzed using statistical package for the social science (SPSS), then was interpreted by using statistical analysis graph, charts and measure of central tendency (mean, median, mode).

### Data analysis plan

**Objective 1:** To assess job satisfaction among HCWs.

**Analysis:** The following variables were included; -job satisfaction, wage satisfaction, benefits satisfaction, advancement satisfaction, supervisor support, coworker support, security, job autonomy, work overload and meaningful work. The research variables used were numerical variables.

Statistical method used was descriptive statistics by assessing frequency of variables in terms percentages. The results were presented in form of tables and bar graphs

**Objective 2:** To determine job relation and benefits among HCWs in health facilities.

**Analysis:** The following variables were included; -supportive work culture, management trust, availability of job benefits, availability of health work programs at work, work to non-work conflict, non-work to work conflict and workplace/schedule flexibility. The research variables used were numerical variables.

Statistical method used was descriptive statistics by assessing frequency of variables in terms percentages. The results were presented in form of tables and bar graphs

## ETHICAL CONSIDERATION

Ethical clearance and permission to conduct this study was obtained from the respective department of research ethics and review committee of CATHOLIC UNIVERSITY OF HEALTH AND ALLIED SCIENCES. Permission was also requested and granted from the district administration of Sengerema in Mwanza region. The aim of the study was made clear to the participants and verbal consent was made with the participants. Confidentiality was observed for the information collected from the participants

## RESULTS

### Social-demographic characteristics of the study participant

A total of 356 health care professionals were included in the study, female participants were more occupying 55.6% while the male were 44.4%. In the study more of the participants were married than those single with percentages of 68.5% and 31.5% respectively. On the aspect of education level most of them had tertiary education level (59%), followed by secondary education (29.8%) and some had primary education (11.2%). Moreover, the cadre which had the greatest proportion in the study was medical attendants (24.7%), followed by registered nurses (20.5%), then 11.8% were clinical officers, and 25.3% included other professionals such as enrolled nurses, assistant medical officers and assistant medical officers as shown on table 1 below.

**Table 1.** Social demographic characteristics

| VARIABLE | FREQUENCY | PERCENTAGE |
| --- | --- | --- |
| **HEALTH FACILITY LEVEL** |  |  |
| Dispensary | 180 | 50.6 |
| Health Center | 176 | 49.4 |
| **GENDER** |  |  |
| Male | 158 | 44.4 |
| Female | 198 | 55.6 |
| **EDUCATION LEVEL** |  |  |
| Primary | 40 | 11.2 |
| Secondary | 106 | 29.8 |
| Tertiary | 210 | 59.0 |

| MARITAL STATUS |  |  |
| --- | --- | --- |
| Married | 244 | 68.5 |
| Single | 112 | 31.5 |
| CADRE |  |  |
| Medical attendant | 88 | 24.7 |
| Registered nurse | 73 | 20.5 |
| Midwife | 33 | 9.3 |
| CO | 42 | 11.8 |
| Lab technician | 30 | 8.4 |
| Others | 90 | 25.3 |

### 4.2 Job satisfaction among health care workers

In this study, participants were required to answer a set of eleven questions which specifically dealt with the state of job satisfaction and participants had to choose a response out of four option and the response were scored and a cut point was made to distinguish between those satisfied and unsatisfied. The results showed that 51.7% were satisfied while a significant 48.3% were not satisfied and this is associated to the fact that 50.6% of the participants were from dispensary level where the working condition is not friendly due to poor transport means and limited resources for medical practice such as absence of laboratory services, inadequate medications and high patient load.

**Table 2.** Job satisfaction among health care workers.

| VARIABLE | FREQUENCY | PERCENTAGE |
| --- | --- | --- |
| JOB SATISFACTION |  |  |
| Satisfied | 184 | 51.7 |
| Not satisfied | 172 | 48.3 |
| EFFECT |  |  |
| Positive effect | 179 | 50.3 |
| Negative effect | 177 | 49.7 |
| WORKPLACE RELATIONSHIP |  |  |
| Positive relationship | 198 | 55.6 |
| Negative relationship | 158 | 44.4 |

### Working environment of health care workers

To assess the working environment workers were required to answer question which reflected the benefits they are required to receive. Workers were assessed whether they were receiving the benefits or not, if they were aware of them and whether they applied to their cadre and setting. In account of some of the benefits 292 of them were beneficiaries of health insurance while 64 were either not beneficiaries, unaware or the benefit does not apply to them, 207 of them were beneficiaries of retirement benefits while 151 were not beneficiaries, unaware or the benefit does not apply to them, there was almost equal distribution of the beneficiaries of paid sick leave with 128 benefiting and 144 not benefiting while 84 were not aware or the benefit does not apply to them, also 132 were beneficiaries of paid vacation days and 148 were not. Moreover, on assistance to education only 74 were beneficiaries and 219 were not while 63 were either unaware or the benefit does not apply to them. This benefit level can be summaried as good thus contributing to a good working environment for health care worker, and this can be reflected to the satisfaction of the workers as shown above.

**Table 3.** BENEFITS

| BENEFITS | YES | NO | DON'T KNOW | DOES NOT APPLY |
| --- | --- | --- | --- | --- |
| Health insurance | 292 | 48 | 10 | 6 |
| Assistance with education | 74 | 219 | 60 | 3 |
| Retirement benefits | 207 | 74 | 68 | 7 |
| Paid sick leave | 128 | 144 | 68 | 16 |
| Paid vacation days | 132 | 148 | 61 | 15 |

### Job satisfaction among workers

Most of the health care workers were at the dispensary level were satisfied at their job (55%) but among the health care workers working at the health center most of them were not satisfied (52%).But among married workers most of them were not satisfied with their job (50.5%) as compared to single workers who were most satisfied (56%).

**Table 4.**
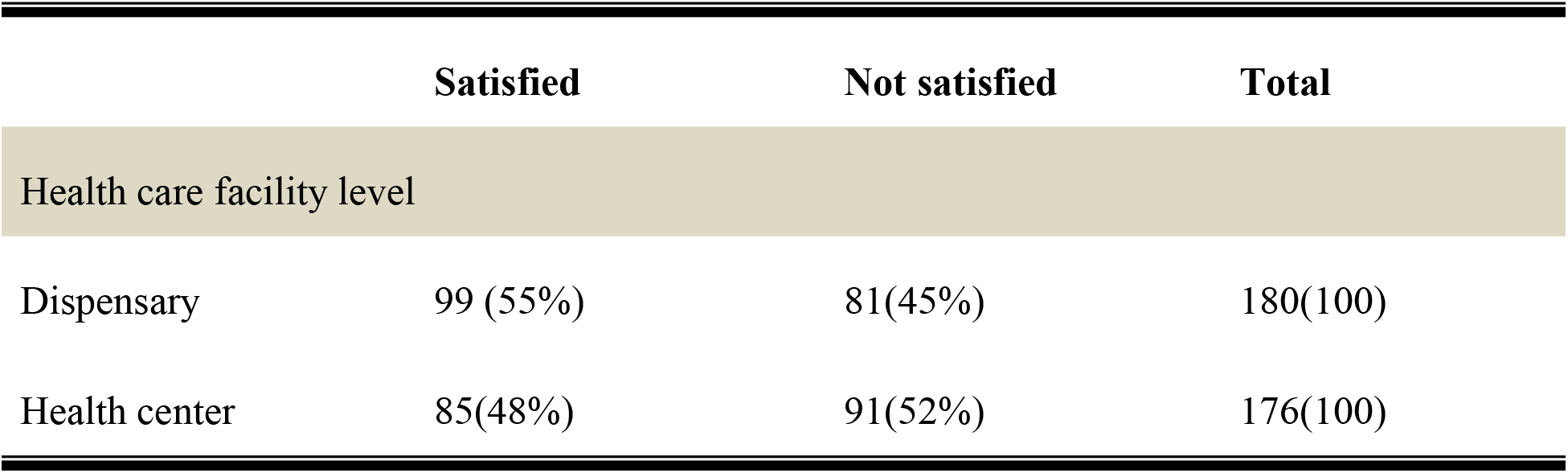
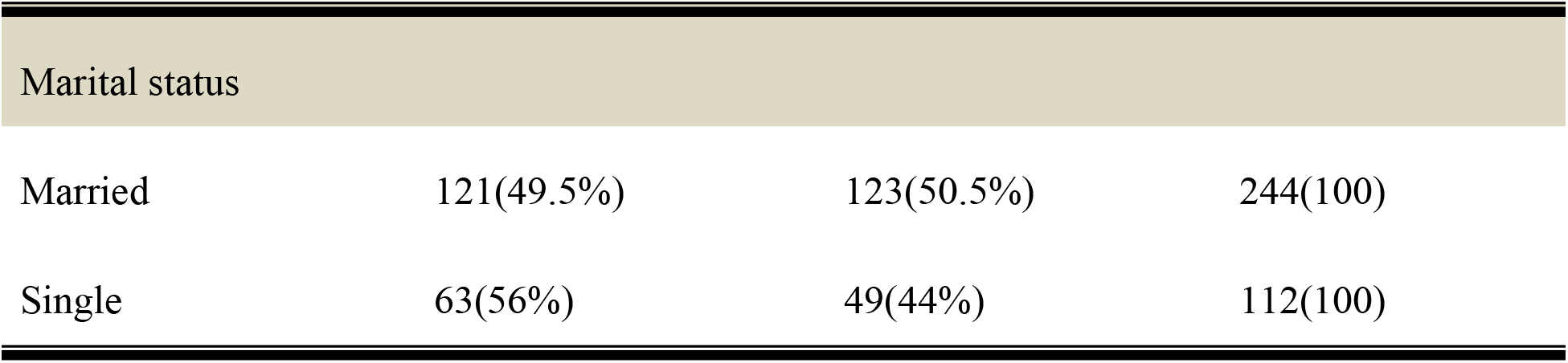
Job satisfaction

## 5.0 DISCUSSION

This study aimed to review the job satisfaction rate of healthcare workers in dispensaries and health centers of Sengerema, Mwanza. Due to the important responsibility healthcare have in prevention, care and treatment, job satisfaction is of high importance. Job dissatisfaction among the staff of such organizations causes an emotional disconnection, apathy and decreased quality of services, which consequently would have serious outcomes.

The overall level of job satisfaction from the total number of participants of health workers in Sengerema health centers and dispensaries was 51.7% for those who were satisfied and 48.3% for those who were dissatisfied in their job. Marital status and professional qualification were found to be the potent predictors of job satisfaction. This finding is supported by other similar studies in Ethiopia in which marital status and type of profession were the predictors of job satisfaction [1, 2]. This result was lower than that found in similar studies conducted in Egypt, Greece, and China, in which the overall level of job satisfaction was 61.24%, 74.63%, and 83.3%, respectively. This difference might be because the economic status and working conditions in these countries are better than those in Sengerema, Tanzania. [3,4,5].

Compared to a study done in West Africa, which was a cross-sectional study conducted on employees of a tertiary hospital in Kano, Nigeria. Where by that majority of the participants were satisfied with their jobs. That is to say, 90.4% of the participants indicated they were satisfied with the jobs they are doing. In this study, even though all categories of health workers were highly satisfied with their jobs; we found no significant association between job satisfaction and the professional categories of the participants.

Another study done in done in Addis Ababa on “Assessment of Job Satisfaction Level and Its Associated Factors among Health Workers in Addis Ababa Health Centers: A Cross-Sectional Study”. Whose results came up as follows: The overall level of job satisfaction was 53.8% with a 95% CI of (48.9%, and 59.0%). These findings appear similar but very slightly higher than our findings, this can suggest similarities in socio-economic and political situations in both of the respective study areas, indicating the need to improve socio-economic status and therefore boost job satisfaction.

This study was also important to identify the factors that affect health professionals’ job satisfaction to focus attention on particular factors and provide possible interventions. Whereby there was a clear association between gender, level of education, marital status with job satisfaction. Results showed that female healthcare workers and healthcare workers with higher level of education had overall higher level of satisfaction than male and health care workers with low level of education. Also, the study has shown that single healthcare workers reported to have higher job satisfaction than their married counterparts. The study also showed the association between working environment and job satisfaction whereby workers in dispensaries had higher job satisfaction than workers who worked in health centers.

## RECOMMENDATION AND CONCLUSION

### RECOMMENDATION

The level of satisfaction and working environment as seen from the study results is good and this is due to presence of benefits to workers and good relationship between workers. Thus, it is important to maintain the good working relationship as well as continue providing the necessary benefits to the workers and those which they are not aware of should be made known to them so as they can be able to enjoy them.

However, do benefits such as education support are not well benefited by the health care workers in the area, thus there should be improvement in the support workers in getting education advancement support from the responsible authority level.

## CONCLUSION

Based on our research, the prevalence of job satisfaction among health care workers in Sengerema district is generally high by 53.9% which shows that most workers are satisfied with their jobs. Most health care workers have good relationship with each other and can depend on each other and they get their benefit as health care workers.

## Data Availability

All data are attached to catholic university of health and allied sciences repository

## LIMITATIONS

1. Some workers requested for things (such as money) in exchange for filling the questionnaire, because they found the research activity as an extra curriculum activity and felt less obliged to participate. This was coped by explaining to the staff of the importance of the research so as to raise awareness of them over the issue, also the situation was later reported to research supervisors so as to avoid this not to reoccur.
2. The research was conducted during work hours thus participation was hard since the workers were simultaneously required to attend patients and provide data for the research, the researchers had to assist them with some works (such as RCH care for women and children coming for clinic) in exchange for the time needed to fill in the questionnaire.
3. Non-medical staffs found it hard to fill the questionnaire due to the difficult vocabularies in it, thus the researchers had to help in translating and filling the questionnaire for them so as to get accurate data.
4. Language barrier; the worker in remote areas found it hard to understand the questionnaire thus it brought difficulties in filling it. This problem was solved by translating the vocabularies to them which was also time-consuming.

## ABREVIATIONS

CUHAS: Catholic University Of heath and Allied Science
H.C.W: Health Care Worker
SAUT: St Augustine University of Tanzania
HIV: Human Immunodeficiency Virus

## Declaration

I, Peter Martin Chilipweli, declare that this dissertation is my own original work and that it has not been presented and will not be presented to any other university. This dissertation is copyright material protected under the Berne Convention, the Copyright Act 1999 and other International and National enactments, in that behalf, on intellectual property.

## Consent for publication

Not applicable

## Availability of data and material

The data sets used and analyzed during the current study are available and still under analysis for subsequent publications but will be available upon request from authors.

## Authors’ contributions

E.K, P M C, A K designed the study, conducted data collection, did data analysis and interpretation of findings, wrote and approved the manuscript. N B provided technical inputs to improve designing the study, supported data analysis, read, improved and E. N Plus D M approved the final manuscript write up.

## Authors Information

P M C is the assistant lecturer at Catholic university of Health and Allied Sciences, (CUHAS) -BUGANDO, Tanzania.

## REFERENCES

1. Tabatabaei S, Mokhber N, Latifian B. Evaluation of job satisfaction among dentists in mashhad. Thequarterly journal of fundamentals of mental health. The Quarterly Journal of Fundamentals of Mental Health. Quarterly Journal of Fundamentals of Mental Health 2004-2005;6:99–104.

2. McManus IC, Keeling A, Paice E. Stress, burnout and doctors’ attitudes to work are determined by personality and learning style: a twelve year longitudinal study of UK medical graduates. BMC Med 2004; 2:29–32.

3. Visser MR, Smets EM, Oort FJ, De Haes HC. Stress, satisfaction and burnout among Dutch medical specialists. CMAJ 2003; 168:271–5.

4. Mesfin Aklilu, Waleleng Warku, Wogayehu Tadele, Yimer Mulugeta, Hussene Usman, Amelework Alemu, Sintayehu Abdela, Alemnesh Hailemariam, Endalkachew Birhanu, “Assessment of Job Satisfaction Level and Its Associated Factors among Health Workers in Addis Ababa Health Centers: A Cross-Sectional Study”, Advances in Public Health, vol. 2020, Article ID 1085029, 6 pages, 2020. https://doi.org/10.1155/2020/1085029

5. Kolo ES. Job satisfaction among healthcare workers in a tertiary center in kano, Northwestern Nigeria. Niger J Basic Clin Sci 2018;15:87–91

6. Deriba, B.K., Sinke, S.O., Ereso, B.M. et al. Health professionals’ job satisfaction and associated factors at public health centers in West Ethiopia. Hum Resour Health 15, 36 (2017). https://doi.org/10.1186/s12960-017-0206-3

7. Bank. W. Discovering the real world, health workers’ career choices and early work experience in Ethiopia. Washington, DC: Africa HumanDevelopment Series; 2010. p. 10.

8. Bass, B. M., & Avolio, B. J., 1992. Multifactor leadership Questionnaire-Short Form 6S. Binghamton, NY: Center for Leadership Studies.

9. Ben-Bakr, K. A., Al-Shammari, I. S., & Jefri, O. A., 1994. Organizational commitment, satisfaction, and turnover in Saudi organizations: a predictive study. Journal of SocioEconomics, 23(4), 449–456.

10. Best, M., & Thurston, N., 2004. Measuring nurse job satisfaction. Journal of Nursing Administration, 34(6), 283–290.

11. Borda, R., & Norman, I., 1997. Factors influencing turnover and absence of nurses: a research review. International Journal of Nursing Studies, 34(6), 385–394.

12. Eisenberger, R., Huntington, R., Hutchison, S., & Sowa, D., 1986. Perceived organizational support. Journal of Applied Psychology, 71(3), 500–507.

13. Elloy, D. F., 2005. The influence of super-leader behaviors on organization commitment, job satisfaction and organization self-esteem in a self-managed work team. Leadership and Organizational Development Journal, 26(2), 120–127.

14. E. T. Bekru, A. Cherie, and A. A. Anjulo, “Job satisfaction and determinant factors among midwives working at health facilities in Addis Ababa city, Ethiopia,” PLoS One, vol. 12, no. 2, Article ID e0172397, 2017.

15. A. Yami, L. Hamza, A. Hassen, C. Jira, and M. Sudhakar, “Job satisfaction and its determinants among health workers in Jimma university specialized hospital, Southwest Ethiopia,” Ethiopian Journal of Health Sciences, vol. 21, no. 1, pp. 19–27, 2011.

16. Aletras VH. Development and preliminary validation of a questionnaire to measure satisfaction with home care in Greece. BMC Health Serv Res. 2010;10:189.

17. Walters BL. Job satisfaction and its modeling among township health center employees in rural China. 2010.

18. Kishk NA, Al Juhani AM. Job satisfaction among primary health care nurses in Al-madinah Al-munawwara. Egypt 2006;81(No.384). https://www.ncbi.nlm.nih.gov/pubmed/17382059.

